# Identifying chest-worn light logger adherence: a validation study

**DOI:** 10.1101/2025.09.29.25336898

**Authors:** Carlyn Patterson Gentile, Adah Thomas, Ryan Shah, Blanca Marquez De Prado, Nichelle Raj, Christina L. Szperka, Geoffrey K. Aguirre

**Affiliations:** Children’s Hospital of Philadelphia, Philadelphia, Pennsylvania, USA; Perelman School of Medicine at the University of Pennsylvania, Philadelphia, Pennsylvania, USA; Sidney Kimmel Medical College at Thomas Jefferson University, Philadelphia, Pennsylvania, USA; Departments of Pediatrics and Neurology, Cincinnati Children’s Hospital Medical Center and University of Cincinnati College of Medicine, Cincinnati, OH, U.S.A

**Keywords:** daily light exposure, continuous light measurements, melanopic equivalent daylight illuminance, photopic illuminance, data reliability

## Abstract

**Background:** Light exposure plays an important role in overall health because it entrains circadian rhythms. Recent technological advances in wearable light loggers allow measurement daily light exposure habits. Using a chest-worn light logger, our goal was to 1) develop methodology for differentiating adherent versus non-adherent use, and 2) define differences in lighting intensity in indoor and outdoor environments, to improve data reliability in future clinical studies using this technology.

**Methods:** Five testers used a 10-channel chest worn light logging device under different conditions of wear and non-wear, and across a variety of indoor and outdoor lighting environments. Measurements from the light logger (photopic illuminance, device orientation, accelerometer data, time of day) were used to train and validate a logistic regression model to differentiate wear from non-wear and correct nighttime placement. This model was then applied to 20 adolescents and young adults with migraine who wore the light logger device for one week. Furthermore, measurements of photopic illuminance and melanopic equivalent daytime illuminance (mEDI) of indoor versus outdoor lighting environments were compared to identify the optimal distinction point between darker indoor and brighter outdoor environments for the chest-worn light logger.

**Results:** Movement, device orientation, light, and time-of-day used as predictors in a logistic regression model had excellent differentiation between wear, non-wear and nighttime use (AUC 0.93 – 0.94), and retained good-to-excellent differentiation when applied to the validation dataset (AUC 0.84 – 0.91). When this model was applied to 20 participants with migraine, we found that 92.9% of participant-days and 71.4% of participant-nights demonstrated at least 80% appropriate use. For differentiating indoor and outdoor lighting environments, the optimal cut-point was 442 lux for photopic illuminance, and 412 lux for mEDI.

**Conclusions:** We demonstrate that internal measurements from a chest-worn light logging device can reliably differentiate wear from non-wear. We also found that the optimal cut-off to differentiate indoor and outdoor lighting environments was similar though slightly lower than then 1,000 lux cut-offs traditionally used to define bright light conditions. These findings can be used to improve data reliability in studies of everyday light exposure in clinical populations using chest-worn light loggers.

## Introduction

Wearable light loggers are a rapidly advancing technology that provide a tool to measure an individuals’ light exposure during daily life. This has important health implications, as reduced light exposure during the day and increased light exposure at night is linked to sleep disruption^1–3^ and early mortality.^4^ Wearable light loggers provide a tool to address this modifiable risk factor to improve health outcomes.

Wearable light loggers are now commercially available that can isolate melanopic equivalent daylight illuminance (mEDI i.e. “blue light”), giving rise to signals important to circadian photoentrainment,^5^ from photopic illuminance that provides the basis for image-forming vision. This capability has great potential to be used in clinical populations to understand how exposure to different light signals impact health and disease. Additionally, design changes allow light loggers to be worn on the chest (i.e. clipped to the chest or as a pendant around the neck), capturing light closer to the visual plane than a wrist worn watch.^6,7^ Demonstrating the clinical relevance of such measurements, we performed an exploratory study of adolescents and young adults with migraine using chest-worn light loggers that showed participants with temporal 24-hour mEDI profiles shifted to later in the day reported more frequent headache.^8^

Given the recency of this technology, factors remain that limit the interpretation of light logging results. First, there is not a standardized approach to confirm when participants are wearing the light logger, constraining data reliability. Second, “bright light” environments have been defined as > 1,000 lux (either photopic illuminance or mEDI) to approximate time spent outdoors (1,000 to 100,000 lux) from indoors (generally < 1,000 lux, depending on lighting conditions),^9–11^ but this distinction has not been directly tested using chest-worn light loggers.

To address these limitations in this pre-registered study,^12^ we describe an approach using the internal sensors within a chest-worn light logger to identify adherent use. We validate this approach and demonstrate its application in our previously published study in adolescents and young adults with migraine.^8^ We hypothesized that the light logger’s orientation sensor, accelerometer, light measurements, and time-of-day would be sufficient to separate wear from non-wear periods. Furthermore, we report on light measurements of indoor and outdoor environments to determine the optimal cut-point for a chest-worn light logger.

## Materials and Methods

This study was pre-registered.^12^

### ActLumus Measurements

ActLumus devices (Condor Instruments, São Paolo, Brazil) are 10-channel wearable light loggers that provide 24-hour continuous collection of photopic illuminance and melanopic equivalent daytime illuminance with an operating range of 1 – 100,000 lux. Data were collected at a sampling rate of 1/minute. We have validated the tabular spectral sensitivity functions of the Light logger, which we previously described.^8^

### Study Design 1: Differentiating Wear from Non-Wear

#### Data collection

Four testers (T1 – T4) collected continuous ActLumus data for 2 – 14 days under the following conditions: wear during active periods, wear during sedentary periods, wear in indoor environments, wear in outdoor environments, nighttime placement with the ActLumus facing up (adherent), nighttime placement with the ActLumus facing down (non-adherent), non-wear periods in stationary places with light (e.g. sitting on a counter), non-wear periods in stationary dark places (e.g. inside a drawer), non-wear periods where the device is in a mobile dark place (e.g. in a pocket or bag). Light diary data were collected through REDCap (Research Electronic Data Capture)^13,14^ hosted by the institution.

#### Differentiating wear from non-wear

Orientation, time above threshold (TAT) of the accelerometer that captured movement, photopic illuminance, and time-of-day from the light logger were utilized to build the model. An orientation of 2 indicated the device was hanging as a pendant important for identifying wear-time, an orientation of 16 indicated the ActLumus was face up, and an orientation of 32 indicated that the ActLumus was face down. We used upright position (orientation = 2) versus not (orientation ≠2) to help identify wear periods. We used face down (orientation = 32) versus not (orientation ≠ 32) to help identify nighttime use with incorrect placement. We did not use face up orientation as a binary variable to allow any non-face-down orientation as adherent during night wear because we reasoned that the light sensor would be able to measure light. Movement was defined as time above threshold (TAT) > 0, which we predicted would help differentiate wear from stationary non-wear and sleep conditions. Photopic illuminance of 1 lux or less was used to define periods of darkness that we predicted could be useful for differentiating wear from stationary and mobile non-wear and nighttime conditions. Time-of-day was used as we predicted it would help differentiate non-wear from nighttime use. Time windows of 1 minute (sampling rate), and 5, 10, 20, and 30 minutes were used to determine the optimal epochs to separate wear from non-wear.

Models were developed using publicly available custom software developed in Matlab (Mathworks®). Logistic regression and support vector machine learning models were compared, and time-of-day, TAT, photopic illuminance, and device orientation were subjected to principal component analysis and used within the model. Receiver operating curves (ROC) were determined and under the curve (AUC), sensitivity, and specificity were calculated. AUC of 0.8 – 0.9 was considered good discrimination, and AUC of 0.9 and above was considered excellent discrimination. Once the final model was determined, eighty percent of data from T1 who took a detailed and precise account of light exposure was used to generate the models. The remaining 20% of data from T1 and all data from T2 – T4 were used to validate the models.

#### Model application, participants with migraine

To demonstrate application of the model, data from adolescents and young adults who were part of a single-center prospective observational exploratory study conducted within the pediatric headache program within the Children’s Hospital of Philadelphia (CHOP) neurology department between October 2024 and March 2025 were analyzed. The protocol received approval from the Institutional Review Board at CHOP and was in accordance with the Declaration of Helsinki. Methods and results from this study have been previously reported.^8^ Participants continuously wore the light logger for 7 days. During sleep, ActLumus was removed while sleeping to avoid wearing a cord around the neck and to prevent the device from being obscured by bedsheets. Participants were instructed to keep the light logger face up on a bedside table to allow for night recording.^1^ They were also instructed to avoid wearing it in the shower or submerging the device in water. Those who participated in an activity that prevented them from wearing the light logger were instructed to keep the device in the same area with the sensor unobscured.

We recalculated light metrics previously described in our exploratory study,^8^ by applying the wear/non-wear model. 24-hr light profiles were visually represented as the mean log-photopic illuminance with 95% confidence intervals calculated by bootstrap analysis. Light intensity was captured as photopic luminous exposure (i.e. the total 24-hour photopic light exposure expressed in kilolux^*^hr) and minutes per day exposed to bright light (photopic illuminance > 1,000 lux). Light timing was defined by percent time spent within recommended mEDI limits for optimal melatonin activity.^15^ Specifically, we determined the percent time spent at a minimum 250 lux mEDI from 7:00 – 17:00, percent time at a maximum of 10 lux 20:00 – 23:00, and 1 lux or less from 0:00 – 6:00.^15^ Fixed epochs were based on the photoperiod and the school schedule. We estimated shifts in the mean 24-hr light profile by calculating the correlation between a shifted version (+/-200 minutes) of each participant’s profile to the mean across participants and taking the highest correlation value to determine the size of the shift, as previously described.^8^

### Study Design 2: Differentiating indoor and outdoor lighting environments

#### Data Collection

Co-author and tester (T5) collected detailed measurements for at least 5-minute periods to compare indoor and outdoor lighting conditions. Outdoor recordings were collected after sunrise and before sunset to capture natural daytime light. Indoor recordings included residential, office, retail, and transportation environments. All observations were recorded during July 2025 in the Philadelphia area or Washington DC. The light logger device was worn as a pendant or placed face-up on a surface during observations.

#### Analysis

Analyses were conducted in R Studio version 4.4.2. LightLogR package^16^ was used to visualize the data and calculate the geometric mean of photopic illuminance and mEDI across indoor and outdoor observations. Above ground transportation was also shown for reference. Youden’s index and receiver operating characteristic (ROC) analysis were then used to determine the optimal cut-points between indoor and outdoor light levels for both photopic illuminance and mEDI. Observations of above ground transportation were excluded from the analysis as they represented environments in between outdoor and indoor but are reported in the figures. Analysis was conducted using published and publicly available pROC, dplyr, and tidyverse R packages.^17–20^

## Results

### Study 1: Identifying wear versus non-wear time

To develop and validate a model that differentiates Light logger wear from non-wear, we utilized data collected from testers in a variety of wear and non-wear conditions with corresponding light diary information. Eighty percent of data from tester T1 was used to develop the model. We did not find that smoothing or binning the data into larger epochs (5, 10, 20, and 30 minutes) improved model performance. Thus, we used the original 1-minute sampling rate for the model input. We also did not find that combining variables through principal component analysis or using a support vector machine classifier improved model prediction, thus logistic regression was used. A schematic of the final model is shown (Figure 1a). TAT from the accelerometer data was used to determine periods of movement versus non-movement, orientation was used to determine if the device was in an upright orientation (i.e. the orientation when it is worn around the neck) and if it was face down, photopic illuminance was used to determine dark from non-dark periods. Movement, upright hanging position, face down position, dark (all binary variables), and time-of-day in minutes (continuous variable) were used as predictors in the wear versus non-wear model. Three possible outcomes were defined: wear during the day, non-wear, and appropriate night placement. We used cut points that optimized model function. The model had excellent differentiation between wear (AUC = 0.94, sensitivity = 0.85, specificity = 0.98), non-wear (AUC = 0.94, sensitivity = 0.88, specificity = 0.93), and nighttime periods (AUC = 0.93, sensitivity = 0.89, specificity = 0.91). The remaining data from testers T1 - T4 were used to validate the model. This model retained excellent differentiation for wear periods (AUC = 0.91, sensitivity = 0.81, specificity = 0.98), and good differentiation for non-wear (AUC = 0.94, sensitivity = 0.64, specificity = 0.84), and nighttime periods (AUC = 0.84, sensitivity = 0.75, specificity = 0.83) when applied to the validation dataset.

**Figure 1:**
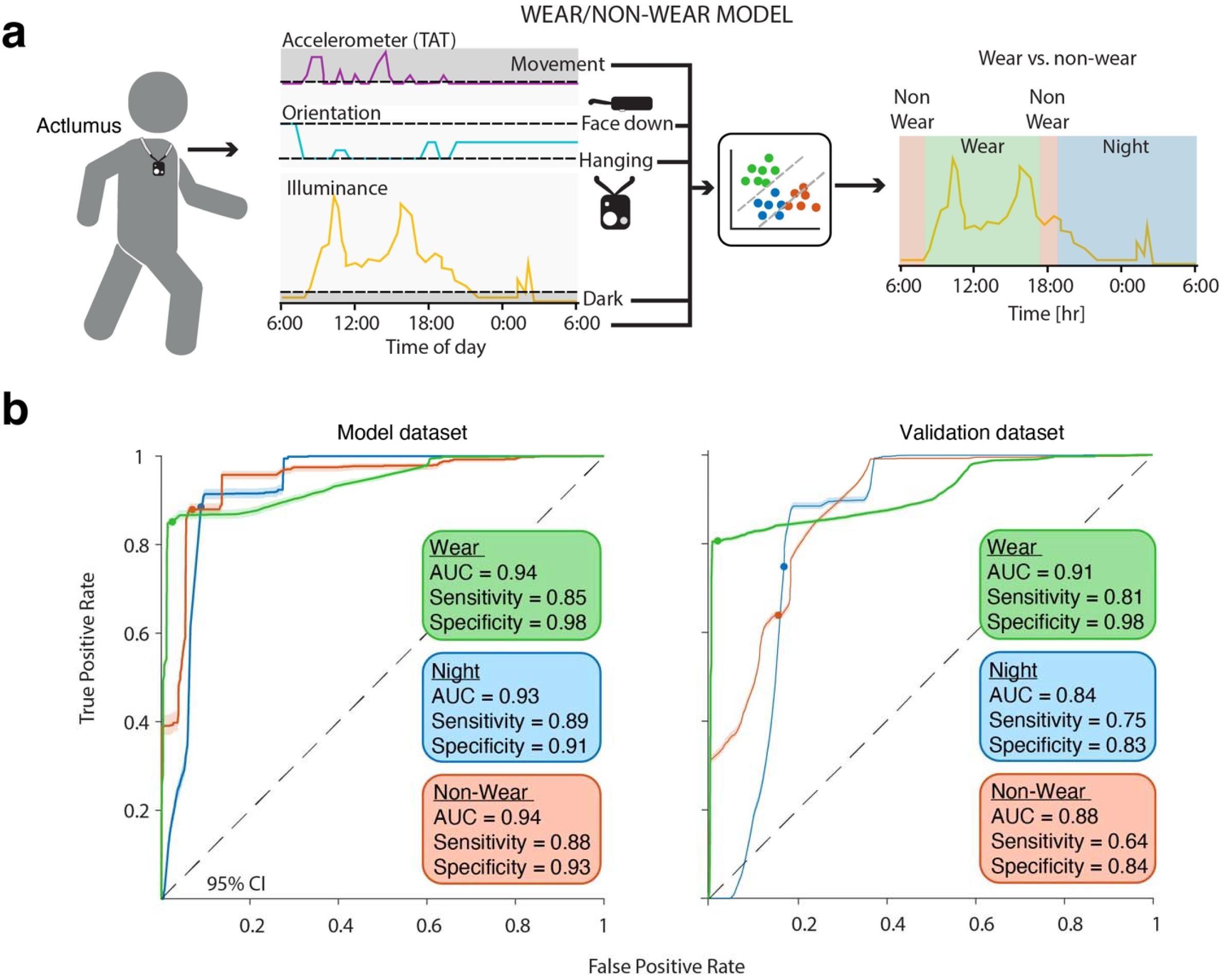
Wear/non-wear model development and validation. a) Schematic of the wear/non-wear model. Measurements of the light logger orientation, accelerometer-based movement, and illuminance were used to determine when the device was hanging in an upright position, face down, when there was movement, and when it was dark. These binary predictors were added to time-of-day to a as logistic regression model of wear, non-wear, and appropriate nighttime use. b) Model development (left) and validation (right) for the three wear categories with AUC, sensitivity, and specificity. Error bars represent 95% confidence intervals.

We then applied the model to 20 participants with migraine who wore the light logger for 1 week, comprising 140 participant-days, whose data has been previously published (Figure 2).^8^ We used the predicted wear and non-wear habits to define adherent daytime and nighttime use. Adherence was defined as at least 80% wear/proper use during a defined time window. We separated adherent day use (10:00 – 20:00) and adherent night use (0:00 – 06:00), comparing wear and night use, respectively, to periods of non-wear. These time frames were chosen to account for differences between weekdays and weekends.^8^ Using these criteria, 92.9% of days and 71.4% of nights were adherent (Figure 2a). There was a substantial reduction in photopic illuminance on non-adherent compared to adherent days, indicating these criteria are capturing periods of time when the device is not exposed to light, consistent with non-wear. There was a significant increase in nighttime light exposure captured on adherent compared to non-adherent nights, consistent with the device being face up versus face down, respectively. This was most dramatic from 20:00 – 24:00, even though this period was not used to define nighttime adherence. This suggests that the parameters can capture periods of time when the device was placed face down for the night.

**Figure 2:**
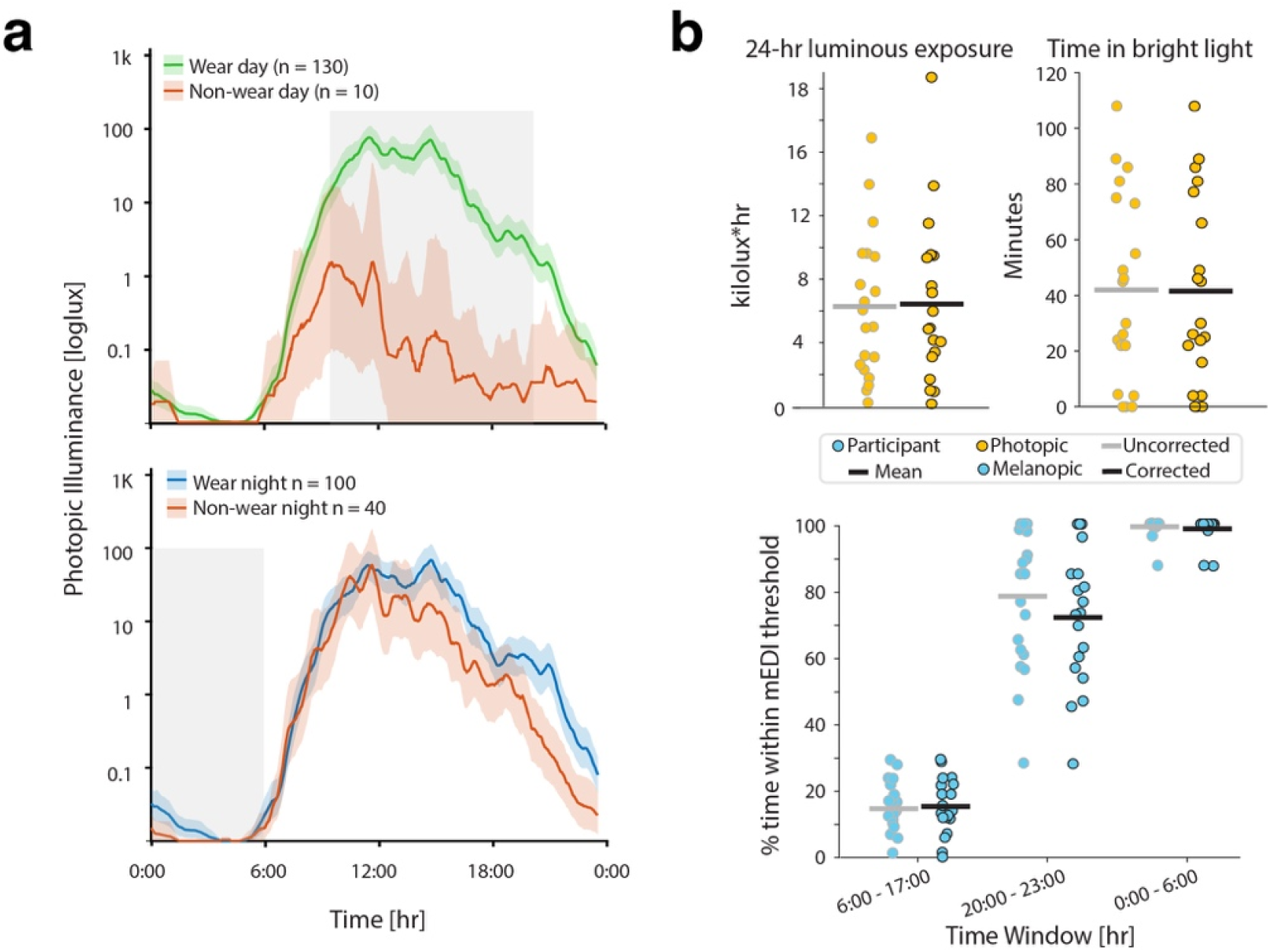
Model application to continuous light logging data from youth with migraine during a typical week. a) Comparison of wear vs. non-wear days (top) and adherent vs. non-adherent nights (bottom). Error bars represent 95% CI by bootstrap analysis. b) Total luminous exposure, time in bright light, and percent time spent within recommended light levels during the day, evening, and night hours based on the original analysis (gray), and after applying the wear/non-wear model (black). Circles represent participant values, and black lines represent the mean.

We then used the adherent data to calculate percent time spent within recommended light exposure levels. One participant did not have any days that met the threshold for adherence and was removed from further analysis. This is largely consistent with what they reported during the study – they responded that they did not wear the device at all for 3 of the days, and the model indicated there was only partial wear on the other days. The remaining participants had 6 days or more of useable data, and 17/19 had at least 3 days of useable night data. Removing days and nights the model determined to be non-wear time did not substantially change the group averages of the intensity or timing of light exposure (Figure 2b). The largest difference was seen in percent time spent under 10 lux mEDI between 20:00 – 23:00: 78.1% from the original study vs. 72.4% when only the adherent data was used. This indicates that when the device is not face down it is able to capture light use before bed more effectively. Finally, the association we observed between light shift and headache (Rho = 0.66, p = 0.002 vs. Rho = 0.50, p = 0.028) and bad headache (Rho = 0.60, p = 0.005 vs. Rho = 0.47, p = 0.044) days per month was weaker but remained significant.

### Study 2: Differentiating indoor from outdoor lighting environments

Sixty-six environments (21 outdoor, 45 indoor) representing a variety of light conditions were captured by Tester 5. One observation with an unusually low lux level was excluded, as the device may have been obscured by clothing or recorded improperly. Outdoor environments ranged from urban and forested areas with clear sky days to completely overcast days with precipitation, and in shaded and exposed areas. Indoor lighting environments included areas with LED, incandescent, and fluorescent lights, artificial light only, natural light only, and a combination, and ranged from dimly to brightly lit. Indoor environments also included smart phones as the main light source. Examples of various lighting environments with respective photopic and mEDI lux are shown across one day of recording (Figure 3a).

**Figure 3:**
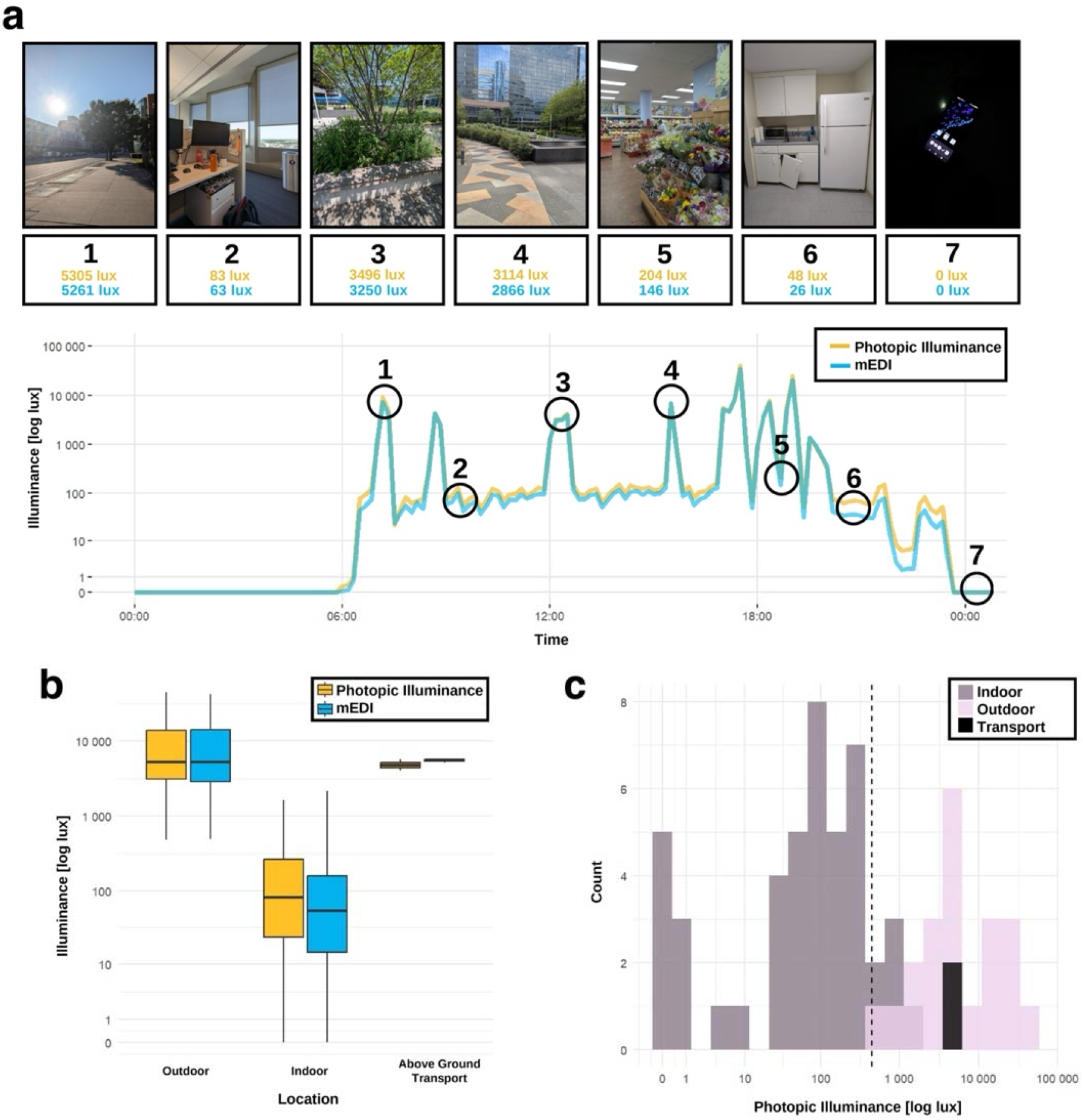
Differentiating indoor and outdoor environments using a chest-worn light logger. a) Example recording day demonstrating the photopic illuminance and mEDI measurements for various indoor and outdoor environments. b) Geometric mean box plot of photopic illuminance and mEDI for outdoor and indoor environments, and above ground transportation. c) Histogram showing the photopic illuminance of each observation with the cut-point for differentiating indoor from outdoor environments. Observations in above ground transport were more consistent with outdoor lighting environments.

The geometric mean of outdoor photopic illuminance was 10,241 lux [range of 484 - 44,427 lux], and the geometric mean of indoor photopic light was 202 lux [range of 0 – 1,652 lux] (Figure 3b). Above ground transportation fell in a similar range to outdoor values. ROC curves were calculated. The cut-point of 442 lux had excellent accuracy for photopic illuminance (AUC = 0.99, sensitivity = 0.89, specificity = 1; Figure 3c), and as did a cut-point of 412 lux for mEDI (AUC = 0.99, specificity = 1, sensitivity = 0.91). We then applied these cut-points to continuous data logged by Testers 1 – 4 (Table 1). The original cut-point of 1,000 lux for photopic illuminance or mEDI had good accuracy, with modest improvements in AUC and specificity for indoor environments using cut-points we derived from multiple indoor and outdoor lighting environment measurements.

**Table 1:** Validation of indoor/outdoor photopic illuminance and mEDI cut-points. AUC = area under the curve.

| Cut-point | AUC | Sensitivity | Specificity |
| --- | --- | --- | --- |
| <u>Photopic Illuminance</u> |  |  |  |
| Indoor (1,000 lux) | 0.83 | 0.94 | 0.73 |
| Indoor (442 lux) | 0.85 | 0.92 | 0.79 |
| <u>mEDI</u> |  |  |  |
| Indoor (1,000 lux) | 0.83 | 0.93 | 0.73 |
| Indoor (412 lux) | 0.86 | 0.92 | 0.80 |

## Discussion

In this study, we developed a model that reliably differentiated periods of wear from non-wear and determined optimal cut-points for distinguishing indoor from outdoor lighting conditions for a chest-worn light logger. This will support reliable data collection and interpretation for future studies that measure the impacts of light exposure on health in clinical populations.

Accuracy of our wear/non-wear model ranged from 0.84 – 0.94 for the model training and validation, comparable to models used to identify non-wear for actigraphy devices.^21^ Our findings are consistent with prior work that has demonstrated the feasibility of detecting non-wear periods in wearable light logger studies. Guidolin and colleagues successfully detected non-wear periods with a near-corneal light logger by having the device placed in a black bag and measuring periods of non-movement.^22^ Others have proposed using periods of low variability in light exposure to detect light logger non-wear, though determined these to be optional in high adherence cohorts.^23^ Our model predicted high adherence (over 90% for day and 70% for night use, respectively), and application of our wear/non-wear model did not substantially change outcomes. However, data from one participant was removed from analysis based on the model output. Days with at least 80% wear time demonstrated significantly higher photopic illuminance as compared to days with less than 80% wear time. Comparing model-derived adherent to non-adherent nights demonstrated significantly higher photopic illuminance in the late evening, which indicates evening light exposure is underestimated on nights when the device was not placed correctly. These findings suggest that it is prudent to apply the wear/non-wear model. It is important to note that trying to differentiate wear from non-wear time may lead to underestimating sedentary time when accelerometer data are used.^24^ Our model was more effective at identifying wear time and was less sensitive at detecting non-wear time, which may help avoid this.

To distinguish indoor and outdoor lighting environments, we found that 442 lux photopic illuminance and 412 lux mEDI were optimal cut-points. While these values are in general agreement with prior work, they are lower than the 1,000 lux cut-point which has been traditionally used to differentiate bright outdoor from dimmer indoor environments.^9–11^ This indicates that for a chest-worn light logger device, a lower definition of “bright light” may better differentiate indoor from outdoor spaces. Further study is needed to determine the generalizability of these results, comparing it to different geographic regions and time of year. Our study has limitations that are important to consider when interpreting the results. First, the model is based on data collected from one tester and performs better for this tester than other testers. However, the model still has good-to-excellent discrimination between wear and non-wear periods when applied to the data of other testers. Furthermore, we only explored logistic regression and support vector machine learning classifier models and there may be different modeling approaches that are more effective at differentiating wear from non-wear. There may also be scenarios where the model is more likely to over or under-estimate wear time, including prolonged periods commuting or lying down during the day (e.g. napping). In these circumstances, it would be helpful to augment the model with participant-reported activities.

## Conclusion

We demonstrate that internal measurements from a chest-worn light logger device can be used reliably to determine wear from non-wear and present a test case where this improved data quality. We also report that cut-point values around 500 lux, compared to 1,000 lux, may modestly improve separation of indoor and outdoor environments when chest-worn light loggers are being used.

## Data Availability

Data collected by the researchers is available for this current study upon request to the authors. Data from the participants with migraine that have been previously published is not available due to legal limitations based on the consent form.

## Acknowledgements

We would like to thank the participants for contributing their time and feedback to this study.

## Abbreviations

AUC: Area under the curve
CHOP: Children’s Hospital of Philadelphia
CI: Confidence interval
mEDI: Melanopic Equivalent Daylight Illuminance
ROC: receiver operating curve
TAT: Time above threshold

